# An adaptive testing strategy for efficient utilization of healthcare resources during an epidemic

**DOI:** 10.1101/2022.01.14.22269232

**Authors:** Sreenath Balakrishnan, Safvan Palathingal

## Abstract

Stringent lockdowns imposed as a preventive measure against rapidly spreading epidemics like COVID-19 adversely affect the economy. They also prolong the duration of the epidemic, making the hardship on the economy two-fold. The extended duration observed in strategies rooted in social distancing and lockdowns is often due to the under-utilization of medical facilities. Even though an under-utilized health care system is preferred over an overwhelmed one, an alternate and optimal strategy is to maintain medical facilities close to their capacity. We show that such a control strategy can be achieved by varying the testing rate and we present an algorithm to calculate the number of tests per day to achieve this. We illustrate the efficacy of our strategy by showing that it reduced the epidemic duration by 40% in comparison to lockdown-based strategies. Our strategy helps in managing the epidemic without high fatalities and the crippling effects of lockdowns.

## Introduction

The COVID-19 pandemic has caused millions of deaths and economic hardships throughout the world. The primary fear during an epidemic is that the number of people requiring medical care would far exceed the available healthcare facilities (Fig. 1a). This scenario would cause high mortality due to the unavailability of basic medical care even when the disease is not fatal. This is especially the case with COVID-19 as most of the infected individuals, close to 90%, have mild-to-moderate symptoms. To prevent fatalities due to paucity of healthcare resources, many countries had imposed lockdowns. The objective of such interventions is to decrease the disease-spreading rate so that the number of infected requiring medical care is always less than the available healthcare resources (Fig. 1a). This reduces fatalities at the cost of prolonging the epidemic and economic hardships. The extended duration is primarily because the available healthcare resources are under-utilized for the most part of the epidemic. They are fully utilised only at the peaks of each wave. We propose an alternative strategy wherein the healthcare capacity is completely utilized during the epidemic (Fig. 1a). Such a strategy reduces the duration of the epidemic without high fatalities.

**Figure 1:**
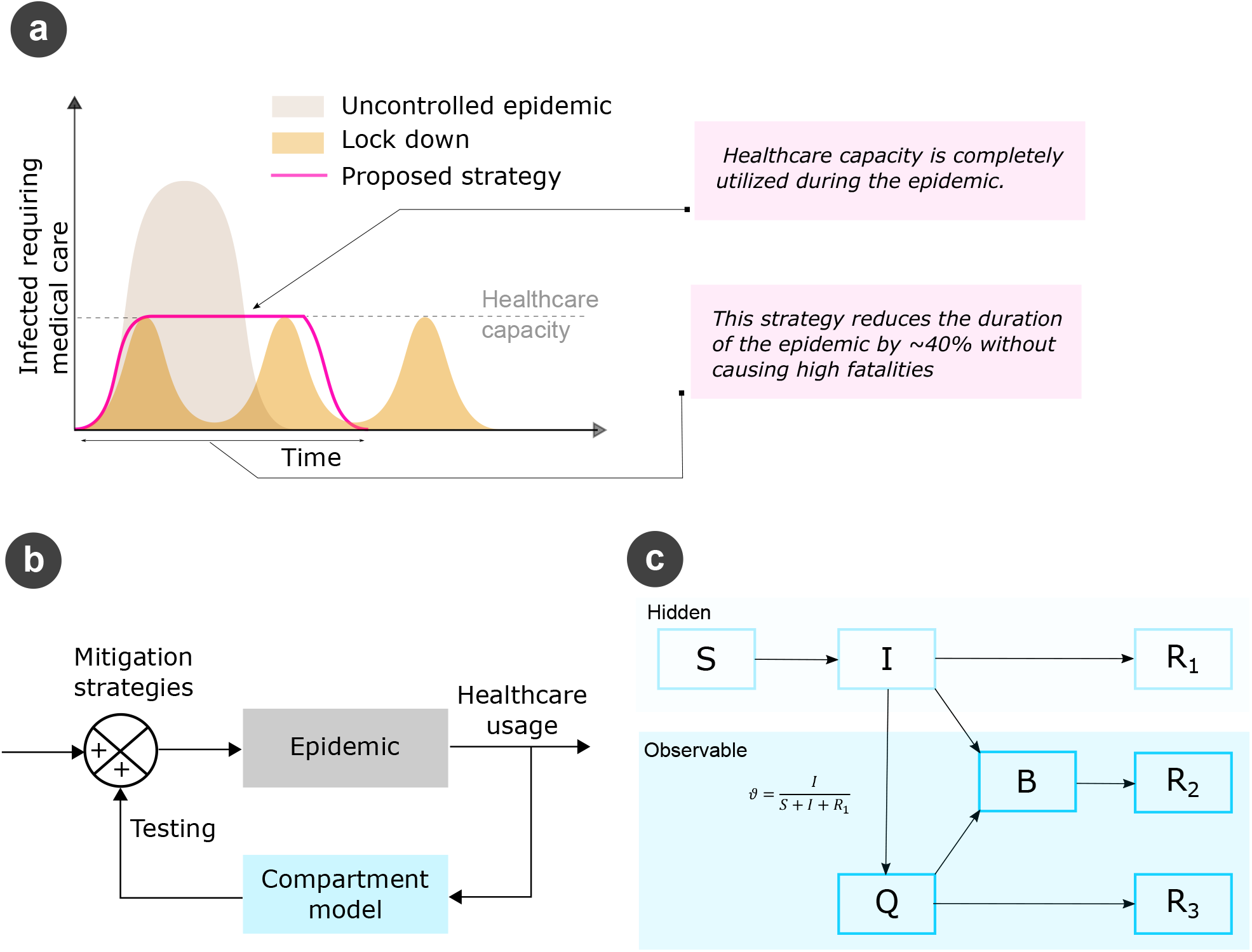
Proposed strategy for efficient utilization of healthcare resources. (a) In an uncontrolled epidemic, the number of infected requiring medical care exceeds the available healthcare facilities leading to high fatalities. Alternating lockdowns and relaxations reduce fatalities by ensuring adequate medical facilities but increase the duration of the epidemic and result in multiple epidemic waves. We propose a strategy wherein the usage of healthcare facilities is maintained at its capacity for reducing the duration of the epidemic while ensuring adequate medical facilities. (b) To realise this strategy, we propose a model-predictive control technique, wherein testing rate (number of tests per day) is used as a control signal. The epidemic is modelled as a simple compartment model (c) with the following compartments; *S* - susceptible, *I* - infectious, *B* - infected requiring the critical healthcare resource, *Q* - quarantined, and *R*_1,2,3_ - recovered from their respective connected compartments. *S, I* and *R*_1_ are individuals outside the medical system and are hence hidden variables. *Q, B, R*_2_, and *R*_3_ are within the medical system and are hence observable variables. By testing random individuals from the hidden population, we identify infectious individuals and quarantine them (*I* -> *Q*) thereby controlling the spread of infection. Test positivity is also an observable variable. The testing rate required to maintain the usage of healthcare facilities at its capacity is expressed in terms of the observable variables and fed back into the epidemic to control it.

The proposed strategy (pink curve in Fig. 1a) requires intricate control of the epidemic, which may not be possible using gross interventions such as lockdowns. We hypothesized that this could be achieved by varying the testing rate. Previous studies with testing and tracing have focused on simulating their effects at various uniform rates and recommending the suitable regime among them (*1–4*). It may be noted that the final recommendation from these studies is a fixed rate of testing or tracing, which typically under-utilise the medical facilities (*3*). In contrast, here we control the epidemic by using testing as a control signal, which is modulated in real-time based on the current occupancy and rate of increase of occupancy of the medical facilities. This method can be adopted by local administrative units such as district, town or county with a sizeable population and associated medical facilities.

We use a model-predictive control technique for realizing this strategy (Fig. 1b). Testing rate, number of tests per day (*p*), is calculated using a simplified compartment model that contains quarantine (*Q*) and critical healthcare resource (*B*) compartments (Fig. 1c). The critical medical resource is the one that gets exhausted first, which could be ICU beds, beds with oxygen facility or ventilators. The occupancy of the compartments within the healthcare system, *B, Q, R*_2_, and *R*_3_, are known and therefore observable variables. The occupancy of the compartments outside the healthcare system, *S, I*, and *R*_1_, are unknown and therefore hidden variables. To control the epidemic, the infectious individuals (*I*) among the hidden population should be identified and quarantined. This is achieved by testing random individuals from the hidden population. Test positivity *ν*, fraction of infectious individuals in the hidden population, is also an observable variable. We derive an expression for *p* required to maintain *B* at *B*_*max*_ in terms of the current values of the observable variables (see methods section). This expression and the values of the observable variables at the end of each day are used to calculate the number of tests for the next day. By using this testing rate, the epidemic progression for the next day is simulated and the process is repeated until the end of the epidemic. Epidemic progression can be adequately simulated only using complicated models such as network or agent-based models. However, to demonstrate the feasibility of our strategy, here we have simulated the epidemic evolution using the same compartment model shown in Fig. 1c.

## Results

As mentioned before, we intend to keep the critical resource at *B*_*max*_ to minimize the duration of the epidemic. In order to smoothly lead *B* from zero to *B*_*max*_ in a manner that facilitates mathematical modelling, we force *B* to trace a sigmoidal curve that increases from zero to *B*_*max*_ and stabilizes there. For converging *B* to the sigmoid, we match 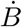 with the slope of the sigmoidal function by defining 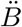 as the difference between them. This desired 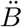, can be achieved by controlling 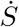, the rate at which the infection is spreading among the susceptible individuals. 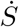 can be modulated by regulating the number of infectious individuals in the hidden population, which in turn can be controlled through testing, *p*. We develop an algorithm to control the epidemic by updating the testing rate based on the observable variables on the previous day.

We first evaluate the performance of our adaptive testing algorithm in controlling the epidemic (Fig. 2a- c). We assume the following parameters for the epidemic: total population *N* = 1.5 million (approximate population of the state of Goa, India); *B*_*max*_ = 0.2 per 1000 (*5*); *R*_0_ = 2.0 (*6*); fraction of infected requiring critical resource, *δ* = 2% (*7*); average duration for recovery from *I* and *Q*, 1*/α*_1_ = 7 days (*1, 7*); and average duration for recovery from *B*, 1*/α*_2_ = 15 days (*7*). The adaptive testing strategy could maintain *B* at *B*_*max*_ throughout the epidemic. The strategy is imposed when *B* increases beyond 20% of *B*_*max*_, corresponding to the point where the colour of the curve changes from brown to pink in Fig. 2a. *B* increases smoothly to *B*_*max*_ and further stays constant at that level (pink curve in Fig. 2a). The epidemic tails off after herd immunity has been achieved (grey curve in Fig. 2a). The corresponding variations in the number of infectious (solid curves) and quarantined (dashed curves) individuals are shown in Fig. 2b. The number of infectious individuals reaches a peak of less than 5 per 1000 just before the epidemic declines. The variation in tests per day required to achieve this is shown in Fig. 2c. When testing commences, there is an instability that rapidly stabilizes within 20 - 30 days. This can be seen from the fluctuations in testing rate shown in the inset in Fig. 2c. These fluctuations are because the testing rate is inversely proportional to the test positivity, *ν*, (refer to Eq. (18) in the Methods Section), which changes rapidly due to quarantining of infectious individuals (dashed curve in Fig. 2c). After stabilisation, the testing rate reaches a maximum and further decreases smoothly to zero. The maximum number of quarantined individuals is around 2.4 per 1000, early in the epidemic, around the time when the testing reaches a maximum after stabilisation (dashed curve in Fig. 2b).

**Figure 2:**
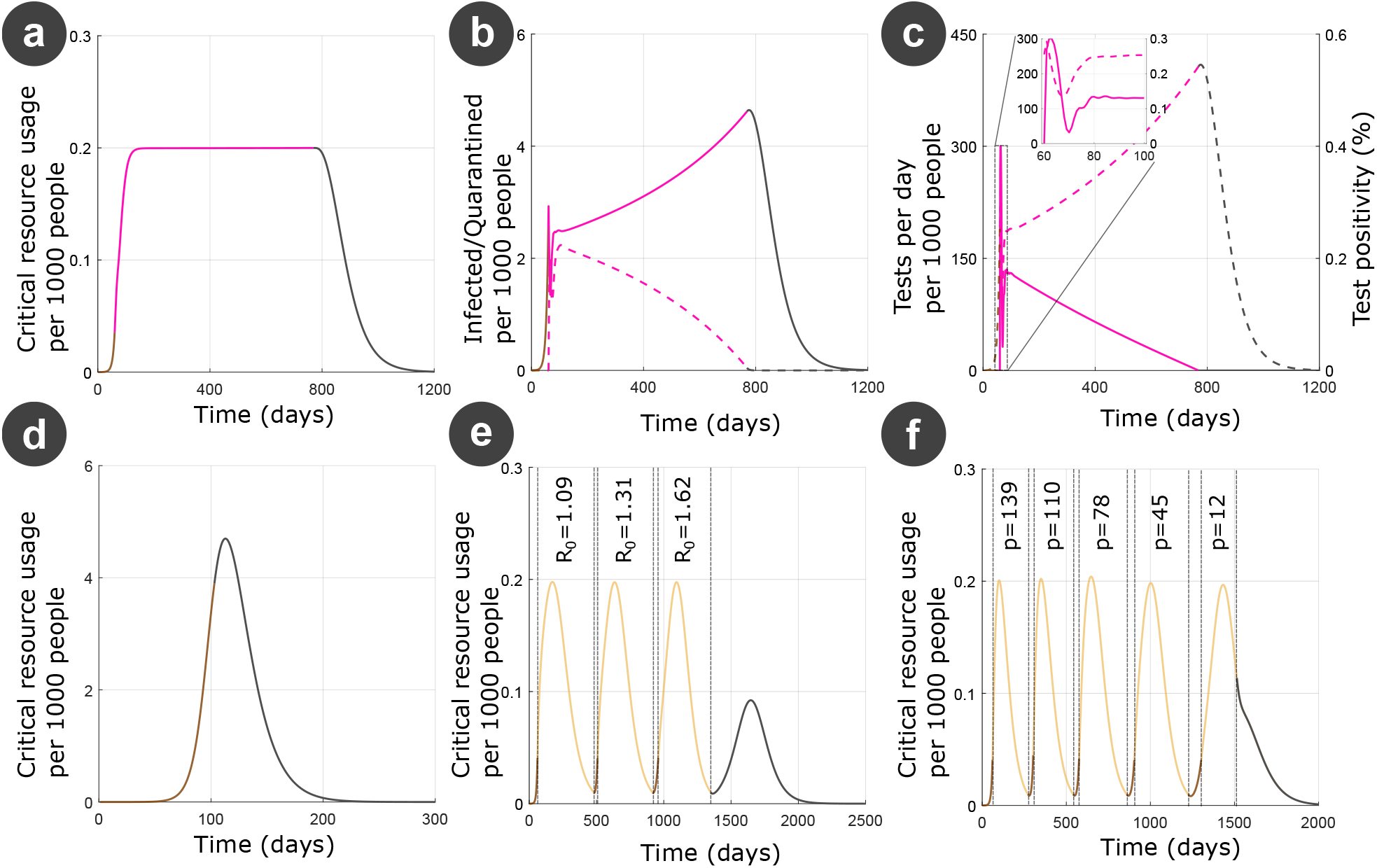
Comparison between epidemic mitigation strategies. Colours indicate different regimes during the epidemic - uncontrolled (brown), mitigation strategies applied (pink in a-c and yellow in e and f), and epidemic winding down (grey) (a-c) Adaptive testing. Evolution of critical resource usage, *B* (a), infected, *I* (solid curves in b), quarantined, *Q* (dashed curves in b), tests per day, *p* (solid curves in c), and test postivity, *ν* (dashed curves in c). (d) Uncontrolled epidemic. Mitigating the epidemic by (e) Controlling *R*_0_ through lock downs or social-distancing norms and (f) constant testing rate.

Unmitigated epidemic, without any interventions, is shown in Fig. 2d. Here, *B* increased to more than 25 times (≈ 5 per 1000 people) its capacity (*B*_*max*_ assumed to be 0.2 per 1000 people), suggesting high fatalities (Fig. 2d). Next, we compare our approach with strategies such as lockdown and constant testing rate (Fig. 2e and f). These mitigation strategies are imposed when *B* goes above 20% of *B*_*max*_ and are discontinued when *B* goes below 5% of *B*_*max*_ (yellow curves in Fig. 2e and f represent the phase when the strategies are active). When the mitigation strategies are relaxed, *B* increases (brown curves in Fig. 2e and f) and the mitigation strategies were reimposed when *B* crossed 20% of *B*_*max*_. This gave rise to multiple waves of the epidemic (Fig. 2e and f). The parameters of the epidemic are same as that assumed for the adaptive testing strategy. In case of lockdowns, *R*_0_ was decreased to ensure that the *B* does not increase beyond *B*_*max*_ in that wave. By restricting *R*_0_ to 1.09, 1.31, and 1.62 for the successive waves, *B* at the peak of each wave was approximately equal to *B*_*max*_ (Fig. 2e). The increasing values of *R*_0_ for successive waves implies easing of lockdown and social-distancing measures as the epidemic progresses. This is tenable because the infection rate for a given number of infectious individuals is lower in successive waves since the number of susceptible individuals decreases as the epidemic progresses. After the third wave, the epidemic winds down (grey curve in Fig. 2e). The total duration of lockdowns (yellow curves in Fig. 2e) is around 1300 days with brief periods of relaxation in between (brown curves in Fig. 2e). Similar results were obtained with a constant testing rate strategy (Fig. 2f). By testing at a constant rate of 139, 110, 78, 45, and 12 per 1000 people per day in five waves, *B* could be restricted below *B*_*max*_ at the peak of each wave. The total duration of this mitigation strategy is around 1500 days.

In comparison to lockdown and constant testing rate strategies, wherein the critical resource is completely utilised only at the peak of each wave, the adaptive testing rate strategy maintains *B* at *B*_*max*_ throughout the epidemic and therefore enables efficient utilisation of healthcare resources. Better utilisation of medical resources reduces the duration of the epidemic. In the adaptive testing strategy, the duration of the epidemic is ≈ 800 days, which is around 40% lower than other strategies. It may be noted that the total number of individuals requiring critical care is approximately the same between these strategies (difference < 10%). This is because herd immunity is achieved after *S* decreases below a threshold, which depends only on *R*_0_ and the number of individuals requiring critical care is assumed to be a fixed percentage of the total infected individuals during the epidemic. Hence, the adaptive testing strategy decreases the duration of the epidemic without higher hospitalisation rates by efficient utilisation of the available healthcare resources.

Furthermore, we tested the performance of our strategy by varying the epidemic parameters such as *B*_*max*_ (Fig. 3 a-c), *R*_0_ (Fig. 3 d-f), *δ* (Fig. 3 g-i), and 1*/α*_2_ (Fig. 3 j-l). In all cases, our algorithm could maintain *B* at *B*_*max*_. Different *B*_*max*_ (0.15 - blue, 0.2 - green, 0.25 - red, and 0.3 per 1000 people - black curves in Fig. 3a-c) correspond to places with different availability of medical facilities. Higher *B*_*max*_ can sustain higher *I* (solid curves in Fig. 3b) thereby achieving herd immunity earlier and consequently reducing the duration of the epidemic (Fig. 3a). Higher *B*_*max*_ does not necessitate higher testing rates. The peak testing rate after stabilisation is equal for different *B*_*max*_ (solid curves in (Fig. 3c) and the duration of epidemic is smaller for larger *B*_*max*_. Different *R*_0_ (1.75 - blue, 2.0 - green, 2.25 - red, and 2.5 - black curves in Fig. 3d-f) correspond to viral strains with different transmissibility. For example, the delta strain of COVID-19 had higher *R*_0_ than previous strains (*8*). Higher *R*_0_ necessitated higher testing rates (Fig. 3f) and the epidemic lasted longer (Fig. 3d and e). The increase in the duration of the epidemic is because the threshold of *S* below which the epidemic declines is lower for higher *R*_0_. Different *δ* (0.5% - blue, 1% - green, 1.5% - red, and 2% - black curves in Fig. 3g-i) correspond to strategies such as vaccinations and isolation of the vulnerable. Alterations in *δ* do not alter testing rates (Fig. 3i) and reduction in *δ* can drastically decrease the duration of the epidemic (Fig. 3g). Different 1*/α*_2_ (11 - blue, 13 - green, 15 - red, 17 days - black curves in Fig. 3j-l) correspond to treatment regimes that can alter the time spent in the critical resource till recovery. For instance, steroids are known to decrease stay in ICUs for COVID-19. Changes in 1*/α*_2_ does not change the peak testing rate (Fig. 3l) whereas reducing 1*/α*_2_ can decrease the duration of the epidemic (Fig. 3j).

**Figure 3:**
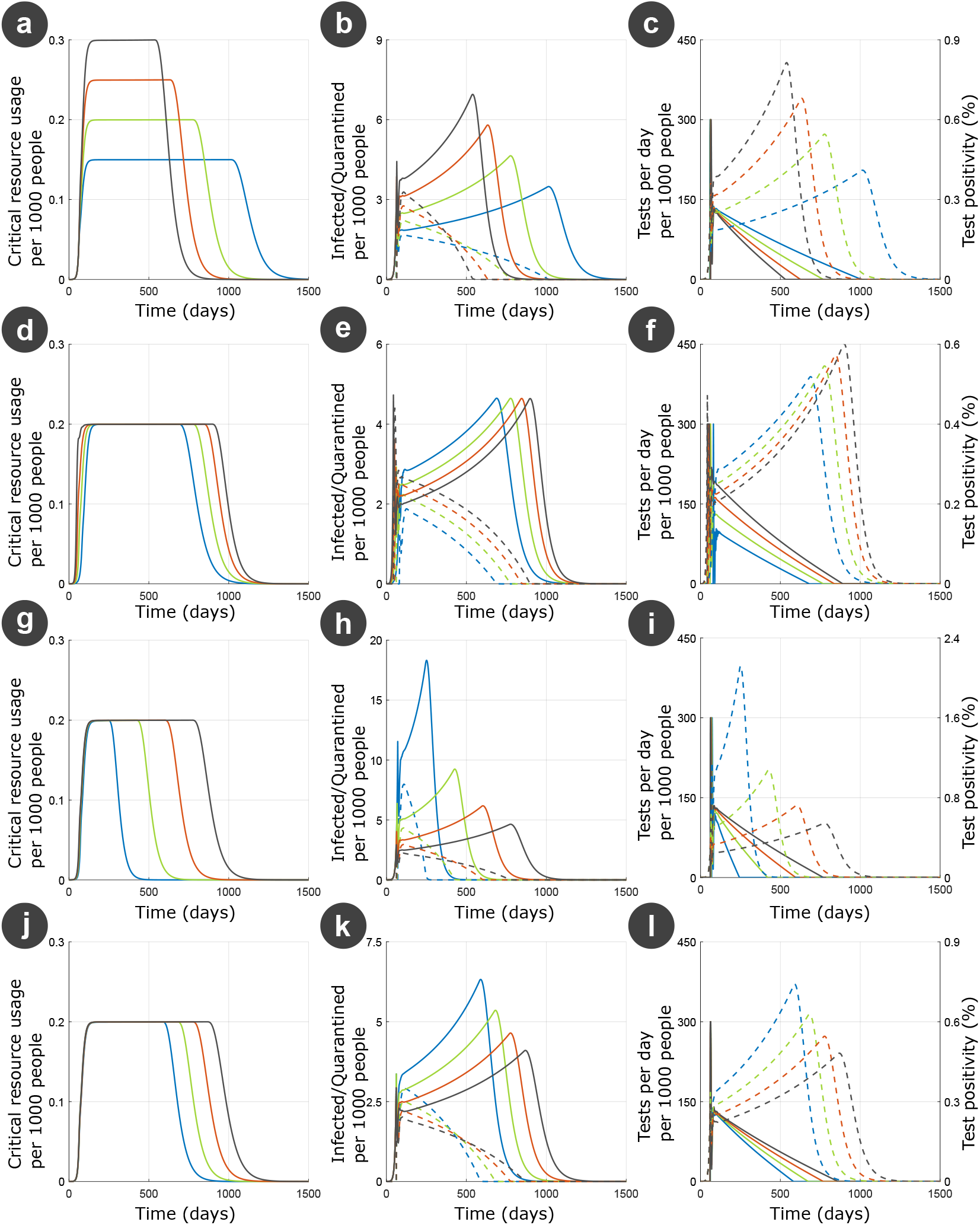
Performance of our strategy for various epidemic parameters. We varied *B*_*max*_ (a-c), *R*_0_ (d-f), fraction of infected requiring critical resource, *δ* (g-i), and average days for recovery in the critical resource, 1*/α*_2_ (j-l). Different values of *B*_*max*_ used were 0.15 (blue), 0.2 (green), 0.25 (red), and 0.3 (black) per 1000 (a-c). Different values of *R*_0_ used were 1.75 (blue), 2.0 (green), 2.25 (red), and 2.5 (black) (d-f). Different values of *δ* used were 0.05% (blue), 1% (green), 1.5% (red), and 2% (black) (g-i). Different values of 1*α*_2_ used were 11 (blue), 13 (green), 15 (red), and 17 (black) days (j-l). When any of these parameters were varied, the other parameters were held constant at the following values *B*_*max*_ = 300, *R*_0_ = 2.0, *δ* = 2%, 1*/α*_2_ = 15. *I* is plotted by solid lines and *Q* by dashed lines in the middle column. Tests per day, *p*, is plotted by solid lines and test positivity, *ν*, by dashed lines in the right column.

## Discussion

In the beginning of the COVID-19 epidemic, a few countries such as UK and Sweden had resisted nationwide lockdowns and tried to achieve herd immunity by allowing the infection to spread through the younger population who are considered to be relatively safe (*9*). However, the rising number of cases and hospitalisations forced these countries to impose strict social-distancing measures (*10, 11*). This was primarily because the healthcare system got stressed and there were no known methods other than social-distancing to blunt the rise in hospitalisations. The method proposed here ‘flattens the hospitalisation curve’, which could have allowed these countries to continue without stringent lockdowns and thereby economic disruptions. In contrast to UK and Sweden, most of the countries imposed strict social-distancing measures until the development of vaccines and drugs to decrease the risk of severe disease and fatalities in high-risk individuals. However, as shown in Fig. 3g and h, even at lower *δ* and 1*/α*_2_ if the epidemic is allowed to spread freely, the medical facilities can get overwhelmed. This too can be avoided by the adaptive testing strategy, which provides an attractive exit strategy without resorting to additional lockdowns.

We showed in Fig. 2 that by flattening the hospitalisation curve, we significantly decrease the duration of the epidemic. For a realistic set of epidemic parameters, chosen based on the available COVID-19 data, we showed that the reduction in duration is 40%. We illustrated the robustness of the strategy by varying the parameters *B*_*max*_, *R*_0_, *δ*, and 1*/α*_2_ (Fig. 3). In other words, for a wide range of values of the epidemic parameters our strategy enables efficient utilisation of medical facilities and thereby decreases the duration of the epidemic.

In practice, three of the epidemic parameters—*B*_*max*_, *δ* and 1*/α*_2_— are controllable unlike *R*_0_ that depends on the transmissiblity of the viral strain. For example, *B*_*max*_ can be improved by building more medical facilities and vaccinations will decrease *δ*. By developing new therapeutic strategies for reducing time spent in hospitals, 1*/α*_2_ can be reduced. The results show that when we combine these measures with the adaptive testing strategy, the epidemic duration can be further decreased. Hence, the adaptive testing is a new paradigm in epidemic control, which incidentally works even effectively in conjunction with the existing pharmaceutical and non-pharmaceutical strategies.

One of the limitations of the method is the very high testing rate required, around 300 per 1000 individuals per day at its peak. In comparison, the peak testing rate in populous countries such as India and US have peaked at around 2-6 per 1000 individuals per day (*12*). Interestingly, a few smaller countries such as Slovakia, Cyprus, and Austria have reported testing rates of 100-150 per 1000 individuals per day suggesting that the high testing rates required by our method are possible (*12*). We could further decrease the testing rates required by our method by combining it with other strategies such as pooled testing, contact tracing and testing random households instead of individuals:

- Pooled testing - Pooling samples can significantly decrease the number of tests required to identify infected individuals in cases of low prevalence (*13*). Many countries such as USA (*14*), Israel (*15*)
- and Germany (*16*) have employed pooled testing during the COVID-19 pandemic. In our case, test positivity < 0.5% (Figs. 2c and 3c,f,i,l), the number of tests can be reduced by a factor of 0.068 (*17*) thereby decreasing the peak testing rate to 20 per 1000 individuals per day. Such testing rates have been reported during the COVID-19 pandemic by several countries such as UK, UAE, Czech Republic, and Denmark (*12*).
- Contact tracing - The fundamental idea of our method is to quarantine enough infected individuals to decrease the infection spreading rate and thereby control the epidemic. In the method developed in
- this paper, all infected individuals are identified by testing random individuals from the population. In such a random sampling strategy, the number of infected individuals identified will be proportional to the fraction of infected in the population, which is very low. The test positivity is of the order of 0.5% (Figs. 2c and 3c,f,i,l), which means that 200 tests are needed to identify one infected individual. In contrast, among a cohort of individuals known to have been in contact with an infected person, the test positivity would be much higher. For example, a recent study concluded transmission probabilities of 4.7% and 10.7% among low-risk and high-risk contacts respectively in two Indian states (*18*), which means 10-24 tests for identifying an infected person among the contacts of another infected individual. Hence, by combining with contact tracing the testing rate can be reduced significantly.
- Testing random households instead of random individuals - Many studies have shown that COVID-19 spreads more rapidly indoors rather than outdoors. Moreover, people do not follow social-distancing norms such as wearing masks in their homes. Hence, when one person in a house gets infected, the chances of the entire household getting infected are very high. Therefore, one individual from a household could be representative of the entire household and if that individual tests positive, the entire household could be quarantined. This is equivalent to redefining the basic infectious unit as a household rather than an individual, which could decrease the number of tests by the average size of a household. Latest UN data shows that average household size of countries varies from 2.2 (France) to 8.8 (Senegal) with India and US at 4.5 and 2.5 individuals per household respectively (*19*). Hence, by testing a representative individual from each randomly chosen households, we could decrease the peak testing rate to 11 - 45%. However, this requires reformulating the compartment model in terms of households, which will also change the contact and recovery parameters.

Social-distancing measures and testing are two distinct non-pharmaceutical techniques used for epidemic control. The former provides gross control over the epidemic. In this work, by using the adaptive testing strategy we showed that the latter gives fine control. We demonstrated this by recasting epidemic mitigation as a control problem wherein the testing rate is modulated in real-time. This is distinct from epidemiology wherein the focus is on predicting the long-term evolution of the epidemic. Epidemiology is extremely challenging because future predictions loose accuracy over time due to error accumulation and emergence of unforeseen circumstances (*20,21*). In contrast, control strategies typically require only short-term prediction. The control signal needs to be updated in a short time window using the most recent data. In our case, we calculate the number of tests for the next day based on the current value of the observable variables. We believe that a simple compartment model is sufficient for such short-term prediction. This is tenable because compartment models can accurately predict epidemic evolution for a couple of weeks. Here, we have used the same compartment model for simulating the evolution of the epidemic, which is a limitation of the current study. In the future, we will simulate the epidemic evolution using complicated models such as network and agent-based models. In such a case, the epidemic parameters of the compartment model that are hidden should be updated in real-time by fitting the model to the latest epidemic data. As mentioned before, *α*_1_, *α*_2_, and *δ* are observable whereas *β* is hidden. *β* could be estimated from the variation in *ν*. We believe that even with complex epidemic evolution models our algorithm can successfully control the epidemic because of adequate short-term predictability of compartment models.

The emerging paradigm for epidemic management is one wherein the government ensures only adequate medical facilities for the infected while delegating the responsibility of protecting oneself from infection to individuals. Our strategy enables such a paradigm without enforcing strict social-distancing measures such as lockdowns. Individuals are required to undergo testing whenever directed by the health authorities and quarantine themselves if tested positive. Hence, our strategy enables efficient epidemic mitigation with minimal intrusion into individual liberty.

## Data Availability

All data produced in the present study are available upon reasonable request to the authors

## Methods

We make the following assumptions for the compartment model. The average number of days for recovery from the *I* and *Q* compartments is 1*/α*_1_ and from *B* is 1*/α*_2_. Quarantined individuals (*Q*) are assumed not to transmit the infection. A fraction of the infected and quarantined, *δ*, are assumed to require the critical resource. The recovered from the *I, B*, and *Q* compartments are *R*_1_, *R*_2_, and *R*_3_ respectively. *p* is the number of tests per day. The governing equations of the model are

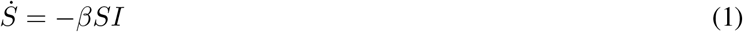

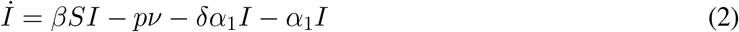

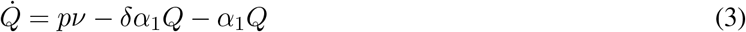

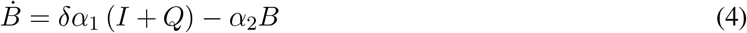

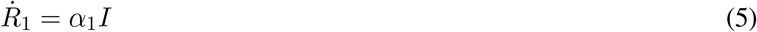

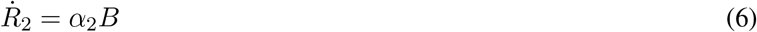

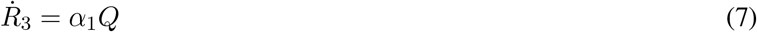

where *ν* is the test positivity given by *I/*(*S* + *I* + *R*_1_) and *δ* (= *R*_2_*/* (*R*_1_ + *R*_3_)) is the ratio of infected individuals requiring critical resource to those who show mild symptoms. It may be noted that only *Q, B, R*_2_ and *R*_3_ compartments and the test positivity, *ν* = *I/*(*S* + *I* + *R*_1_), are observable (Fig. 1c). *S, I* and *R*_1_ are hidden (Fig. 1c). Random individuals from the hidden population, *S* + *I* + *R*_1_ are tested. If *p* such tests are done a day, the expected number of positives is *p* times the proportion of infectious individuals in the total unknown population, *I/*(*S* + *I* + *R*_1_), who are then quarantined.

Our strategy is to force *B* to trace a desired function using testing. The desired function was chosen as a sigmoid that starts at zero and saturates at *B*_*max*_.

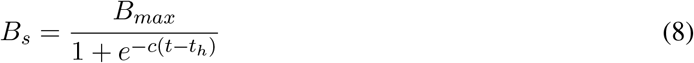

where *c* controls the rise of the sigmoid. For *B* to trace 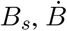 at a given value of *B* should be equal to 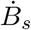, which is given by

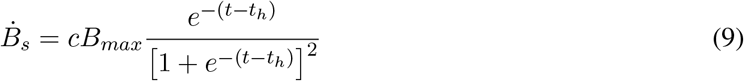

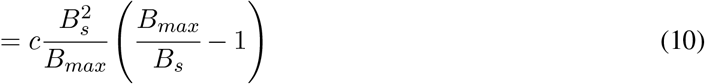

To converge 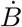 to 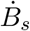, we define a desired 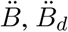, as the difference between them

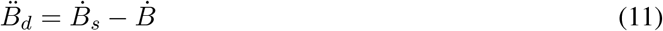

where the time for converging 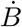 to 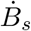 is assumed to be one day. Expression for 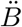 can be obtained by differentiating the expression for 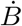 in the compartment model, Eq. (4)

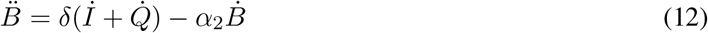

Substituting for *İ*and 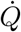 by using Eqs. (2) and (3),

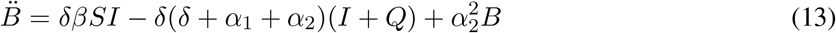

Substituting the desired 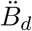 for 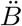 and re-arranging we get the desired infection rate,

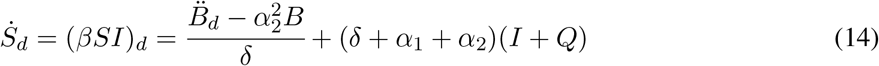

By achieving this desired infection rate, 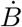 converges to 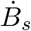 thereby tracing the sigmoid. The desired rate of change of infection rate,

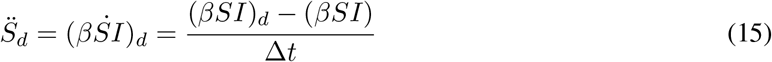

where *βSI* is the current rate of infection and Δ*t* is a time period chosen for smooth convergence of current infection rate 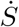 to desired infection rate 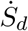 Rate of change of infection rate can be written as

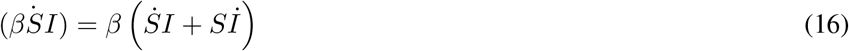

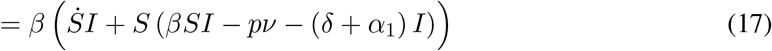

Substituting for 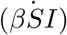 from Eq. (15) and re-arranging

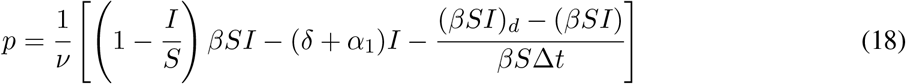

By testing at the rate given by the above equation, the infection rate 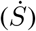 converges to the desired infection rate 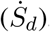, which in turn converges the rate of increase of critical resource usage 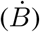 to the gradient of the sigmoid 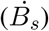. This equation cannot applied directly because it contains hidden parameters such as S and I. For expressing *p* in terms of observable parameters, we use test positivity.

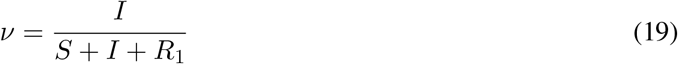

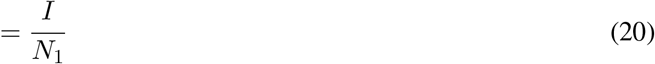

where *N*_1_ = *S* + *I* + *R*_1_ is the number of people in the hidden compartments. By assuming that the total population does not change during the epidemic,

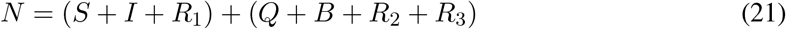

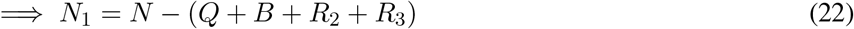

Hence the total number of individuals in the hidden compartments can be calculated from observable parameters and the total population. It may be noted that the people who die due to infection are counted in the recovered compartments. By differentiating the expression for test positivity, Eq. (20),

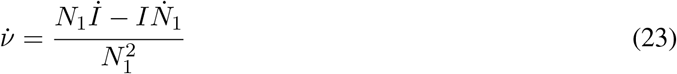

Substituting for *İ*from the compartment model, Eq. (2), and re-arranging we get an expression for the infection rate,

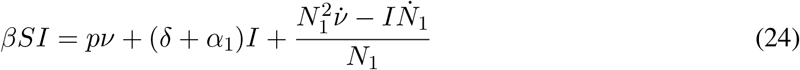

*I* in these equations can be obtained from test positivity as *I* = *νN*_1_ which can then be used to estimate *S*. We have implemented these relations as follows. Testing commences only after a specific percentage of *B* is occupied. For our simulations we have used 20% of *B* as the threshold for starting testing. At the end of each day, *n*, we compute the number of tests required to be performed on the next day, *n* + 1, using the above relations. We first estimate the desired rate of change of 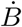 at *n*^*th*^ day 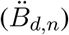 using Eq. (11)

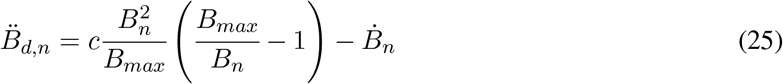

where *B*_*n*_ is the usage of critical resource on the *n*^*th*^ day and 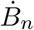 is the rate of change of critical resource usage.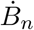 was obtained by calculating the slope of a straight line fit to *B* for the previous three days. Next, we calculate the desired infection rate using Eq. (14)

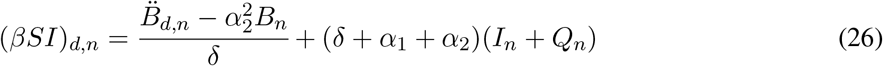

This desired infection rate is used to calculate the number of tests for the *n* + 1^*th*^ day using Eq. (18)

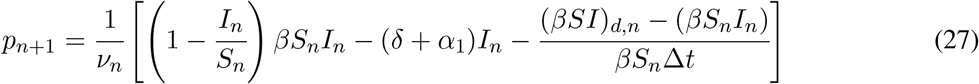

where Δ*t* = 10 days, *I*_*n*_ = *ν*_*n*_*N*_1,*n*_ and *S*_*n*_ is obtained from Eq. (24)

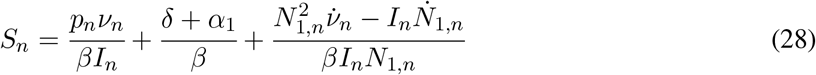

By using this equation, at the end of each day, the number of tests for the next day was updated. This updated testing rate was used to simulate the epidemic for the next day and the process was repeated until the end of the epidemic. Minimum and maximum tests per day was set at 0 and 300 per 1000 respectively to avoid wild fluctuations in testing. The algorithm was implemented in MATLAB (Supplementary Materials).

This strategy was evaluated for epidemic parameters corresponding to the first wave of COVID-19 pandemic in India. The total population of the state of Goa was considered, *N* = 1.5 million. The critical resource was assumed to be ICU beds and its availability was obtained from an official government website, *B*_*max*_ = 0.2 per 1000 (*5*). Reproduction number, *R*_0_ = *β/*(*Nα*_1_), was assumed to be 2.0 (*6*). 1*/α*_1_ = 7 days and 1*/α*_2_ = 15 days were chosen from the recovery periods for asymptomatic and severely infected individuals reported in (*7, 22*). The cumulative branching ratio to severe infection, after accounting for asymptomatic and mild infections, averaged over all age groups was ≈ 0.017 (*1, 7*). This was rounded off to obtain *δ* = 0.02 (2%).

## Acknowledgments

The authors thank Midhun S. Menon for useful discussions, Namrata M. Nilavar for helpful comments, and S.B. thanks IIT Goa for funding (grant number - 2020/IP/SB/008).

