## Supplementary material for "An adaptive testing strategy for efficient utilization of healthcare resources during an epidemic": adaptive_testing.m

```
clearvars;  clc;
close all;

N=1.5e6; alph1=1/7; B_days=15; dlta_f=0.02; p=0; R0=2.0; Bmx=300; %1 wave
dlta=dlta_f*alph1; alph2=1/B_days;
B_fctr=0.2; %fraction of bed usage at which the control kicks in
sig_slp=0.08; %slope of the sigmoidal function
cfctr=0.1; %convergence factor - how close you want to follow the B sigmoidal
pmx=0.3*N; %maximum tests per day
bta=R0*alph1/N;

tend=2000; tv=1:tend;
epi=zeros(length(tv)+1,7); tst_flg=false;
nu = zeros(1,length(tv)+1); N1 = zeros(1,length(tv)+1);
pv = zeros(1,length(tv)+1);
epi(1,:) = [N-1; 1; zeros(5,1)]; 
nu(1)=1/N; N1(1)=N;

for nt=tv
    p = pv(nt);
    h = @(t,x)decay_model(t,x,p,bta,alph1,alph2,dlta);
    [~,xint] = ode45(h,[nt-1 nt],epi(nt,:));
    epi(nt+1,:) = xint(end,:); Q=epi(nt+1,3); B=epi(nt+1,4);
    nu(nt+1) = epi(nt+1,2)/sum(epi(nt+1,[1 2 5]));
    N1(nt+1) = N - sum(epi(nt+1,[3 4 6 7]));
    
    if nt>4
    ply = polyfit(1:2,epi(nt:nt+1,4)',1); dBdt = ply(1);
    dBdt_d = sig_slp*((B^2)/Bmx)*(Bmx/B-1);
    d2Bdt2 = (dBdt_d - dBdt);
        if B > Bmx*B_fctr || tst_flg==true
            tst_flg=true;
            ply = polyfit(1:3,nu(nt-1:nt+1),1); dnudt = ply(1);
            ply = polyfit(1:3,N1(nt-1:nt+1),1); dN1dt = ply(1);

            I_est = nu(nt+1)*N1(nt+1);
            bSI_est = p*nu(nt+1) + (dlta + alph1)*I_est + (N1(nt+1)^2*dnudt - I_est*dN1dt)/N1(nt+1);
            S_est = bSI_est/(bta*I_est);
            bSI_d = (d2Bdt2 - alph2^2*B)/dlta + (dlta+alph1+alph2)*(I_est+Q);
            gma = (bSI_d - bSI_est)*cfctr; %desired change in beta*S*I
            pv(nt+1) = ((I_est/S_est - 1)*(-bSI_est) - (dlta + alph1)*I_est - gma/(bta*S_est))/nu(nt+1);
            pv(nt+1) = round(pv(nt+1));
            if pv(nt+1)>pmx
                pv(nt+1)=pmx;
            elseif pv(nt+1)<0
                pv(nt+1)=0;
            end
        end
    end
    fprintf('time step %d, tests per day %.2f\n',nt,pv(nt+1));
end

figure; hold on;
yyaxis right;
plot(pv*1000/N,'linewidth',2);

set(gca,'fontsize',12); grid on;
xlabel('Time (days)');
ylabel('Tests per 1000');
set(gca,'xlim',[0 tend],'ylim',[0 500],'ytick',0:100:500);

yyaxis left;
plot(epi(:,4),'linewidth',2);
set(gca,'fontsize',12); grid on;
ylabel('Critical resource usage');
set(gca,'ylim',[0 4*Bmx/3],'ytick',0:Bmx/3:4*Bmx/3);
lgnd=legend('Critical resource','Tests');
set(lgnd,'fontsize',12,'location','northeast');
```
