## Supplementary material for "An adaptive testing strategy for efficient utilization of healthcare resources during an epidemic": decay_model.m

```
function dxdt = decay_model(t,x,p,bta,alph1,alph2,dlta)
%DECAY_MODEL SIQBR model in a decay formulation

S=x(1); I=x(2); Q=x(3); B=x(4); R1=x(5); R2=x(6); R3=x(7);
nu = I/(S+I+R1);

dxdt=zeros(7,1);
dxdt(1) = -bta*S*I; %Sdot
dxdt(2) =  bta*S*I - p*nu - (dlta + alph1)*I; %Idot
dxdt(3) =  p*nu - (dlta + alph1)*Q; %Qdot
dxdt(4) =  dlta*(I + Q) - alph2*B; %Bdot
dxdt(5) =  alph1*I; %R1dot
dxdt(6) =  alph1*Q; %R2dot
dxdt(7) =  alph2*B; %R3dot

end
```
